# Population homogeneity for the antibody response to COVID-19 BNT162b2 / Comirnaty vaccine is only reached after the second dose, across all adult age ranges

**DOI:** 10.1101/2021.03.19.21253680

**Authors:** João F. Viana, Marie-Louise Bergman, Lígia A. Gonçalves, Nádia Duarte, Teresa Penha Coutinho, Patrícia C. Borges, Christian Diwo, Rute Castro, Paula Matoso, Vanessa Malheiro, Ana Brennand, Lindsay Kosack, Onome Akpogheneta, João M. Figueira, Conceição Cardoso, Ana M. Casaca, Paula M. Alves, Telmo Nunes, Carlos Penha-Gonçalves, Jocelyne Demengeot

**Affiliations:** CHLO, Centro Hospitalar de Lisboa Ocidental, Serviço de Patologia Clínica, Lisbon, 1449-005, Portugal; IGC, Instituto Gulbenkian de Ciência, Oeiras, 2780-156, Portugal; CIISA, Centre for Interdisciplinary Research in Animal Health, Faculty of Veterinary Medicine, University of Lisbon, Lisbon,1300-477, Portugal; IBET, Instituto de Biologia Experimental e Tecnológica, Oeiras, 2780-901, Portugal; ITQB NOVA, Instituto de Tecnológia Química e Biológica António Xavier, Universidade Nova de Lisboa, Oeiras, 2780-157, Portugal

## Abstract

**BACKGROUND:** While mRNA vaccines authorized for emergency use are administrated worldwide in an effort to contain the COVID-19 pandemic, little is known about the heterogeneity of the humoral immune response they induce at the population scale.

**METHODS:** We conducted a prospective observational longitudinal study in 1245 hospital care workers and 146 nursing home residents, together covering adult ages from 19 to 99 years. Blood samples were taken before vaccination, 3-5 weeks after the first vaccine dose, and 3 weeks after the second dose of BNT162b2 mRNA COVID-19 (Comirnaty, Pfizer/BioNTech). End points were seroconversion to SARS-CoV-2 spike protein and amount of spike-specific IgG, IgM and IgA following the first dose and the boosting effect of the second vaccination. We also addressed the effect of age, sex and pre-exposure to SARSCov-2 on these parameters.

**RESULTS:** Among 1067 baseline seronegative participants, seroconversion after the first vaccine dose varied from a maximum of 98,6% in the younger age strata [19-29 years] to a minimum of 25.4 % in the older age group [70-100 years], while sex had little effect. Large inter-individual variation in the amplitude of the antibody response were observed in all age strata. The second dose brought seroconversion to high frequency (100% and 94.9% in younger and older age strata, respectively) and homogenised IgG responses to high levels, while IgM and IgA levels remained low and heterogeneous. Previous exposure to SARS-Cov-2 boosted IgG level after a single injection. Seven non-responders were identified after the second dose.

**CONCLUSIONS:** In this large study, covering all adult age ranges, BNT162b2 vaccination resulted in isotypic and heterogeneous antibody responses, advocating for the interval between the two doses not to be extended and for serological monitoring of elderly and immunosuppressed vaccinees.

## INTRODUCTION

Authorization for emergency use of two mRNA COVID-19 vaccines, both encoding the most immunogenic protein of SARS-CoV-2, spike, was conceded in late 2020 by regulatory agencies such as FDA and EMA. These authorizations were based on results of phase 3 clinical trials that demonstrated high standards of safety and high levels of efficacy in preventing symptomatic SARS-CoV-2 infections^1,2^. While these vaccines are introduced around the world and administered to millions of people, there is a growing and acute need to evaluate their effectiveness at the population level, an endeavour that may require months of epidemiological studies. Insufficient attention has been given to whether immune responses triggered by mRNA vaccines encoding SARS-CoV-2-spike are homogenously robust. Age and gender are expected factors of variability. Immune responses deteriorate with age, underlying the increased burden of infectious disease, including COVID-19^3^, in older people as well as impaired responses to vaccine challenge^4^. Sex differences have been described in immunity to multiple vaccines in both children and adults, and antibody responses to vaccines are frequently higher in females than males^5^.

To date, humoral immune responses have been seldom measured upon mRNA COVID-19 administration, and when this was the case, limited to the IgG class and concerning rather small groups of participants, ranging from n=8 to 51^2,6-14^.

Immunogenicity of mRNA COVID-19 vaccines and their inter-individual variation can be easily monitored in medium to large cohorts by measuring serum reactivities to the vaccine antigen or part of it. Notably, the receptor binding domain (RBD) of spike contains the amino-acids motifs permitting SARS-CoV-2 binding to the angiotensin-converting enzyme 2 (ACE2) receptor, a prerequisite for infection, and serum reactivity to this region encompasses neutralizing activity^15^. Anti-spike immunoglobulins are also expected to mediate viral particle removal through antibody-mediated opsonization and phagocytosis, and through the recruitment of the complement system. Beyond their direct functionality, vaccine specific antibodies are markers of adaptive immunity responses^16,17^.

The COVID-19 vaccination campaign in Portugal was initiated in late December 2020 coinciding with a peak of disease transmission which reached 131 new daily cases per 100,000 inhabitants and caused an unprecedented demand for hospital care. The vaccination roll-out started with hospital healthcare professionals at the COVID-19 response frontline, soon followed by residents in nursing homes. Here, we report on the humoral response to BNT162b2 mRNA COVID-19 (Comirnaty, Pfizer/BioNTech) vaccination in healthcare professionals and in nursing home residents.

## METHODS

### RECRUITMENT AND ENROLLMENT

The two-cohorts study enrolled 1245 healthcare workers (HCW cohort) from three hospitals, administratively grouped in a single regional centre (CHLO), in Lisbon, Portugal, and 146 residents at four nursing homes (NHR cohort) in Almeirim, a town located in the vicinity of Lisbon, Portugal. All participants enrolled through volunteer sampling. Participants were scheduled to initiate BNT162b2 mRNA (Pfizer/BioNTech, Comirnaty) vaccination along the original protocol of 2 doses with a 3 weeks interval. The study was approved by the Ethics committees of the Centro Hospitalar Lisboa Ocidental and the Administração Regional de Lisboa e Vale do Tejo, in compliance with the Declaration of Helsinki, and follows international and national guidelines for health data protection. All participants provided informed consent to take part in the study.

### BLOOD SAMPLES PROCESSING AND STORAGE

Venous blood was collected by standard phlebotomy. Blood collection occurred at the day of the 1^st^ vaccination (baseline, t0), the day of the 2^nd^ vaccination (3 weeks later for all HCW and 96/146 NHR, and 4-5 weeks later for 50/146 NHR, t1) and 3 weeks after the 2^nd^ vaccine dose (t2). Serum was prepared using standard methodology.

### IMMUNOASSAYS

Electro-chemiluminescence immunoassay (ECLIA) was used to quantify Ig anti-N (Elecsys® Anti-SARS-CoV2 N, Roche) and anti-RBD (Elecsys® Anti-SARS-CoV2 S, Roche), ran (on cobas e602) and analysed as per the manufacturer instructions, with a threshold defining positivity at index value = 0.8 U/ml. Direct ELISA was used to quantify IgG, IgM and IgA anti-full-length spike. The assay was adapted from Amanat et al.^18^ and semi-automized to measure IgM, IgG and IgA in 384-well format, according to a protocol to be detailed elsewhere. Assay performance was determined by testing 1000 pre-pandemic sera and 40 COVID-19 patients diagnosed at least 10 days prior to sera collection. ROC curve analysis determined a specificity of 99.3%, 99.2%, 99.2%, and a sensitivity of 95.9%, 61.2% and 73.7% for IgG, IgM and IgA, respectively. The threshold defining positivity correspond to normalised OD (ODnorm) =1. Serial titration of 67 COVID-19 patients established the assay has a dynamic range of 3 logs titre.

### STATISTICAL ANALYSES

Statistical analyses were carried out using established R scripts. For repeated samples, Quade test was performed to test for differences between strata and Wilcoxon signed-rank test for pairwise comparison of groups (**Figure 2** and **5**). For independent samples, the Kruskal Wallis test was performed to test for differences between strata, and Wilcoxon rank sum test for pairwise group comparison with p-value adjustment for multiple testing (**Figure 3** and **4**).

## RESULTS

### PARTICIPANTS

The study followed 1245 healthcare workers (HCW cohort) and 146 nursing home residents (NHR cohort) vaccinated with BNT162b2 mRNA (Comirnaty, Pfizer/BioNTech) (**Figure 1**). Prior COVID-19 diagnosis was an exclusion criterion, in accordance with the national vaccination plan, and reported cases were restricted to the HCW cohort. Venous blood was collected at the day of 1^st^ vaccine dose administration (time 0, t0), 3 to 5 weeks later at the day of the 2^nd^ injection (t1), and 3 weeks after the 2^nd^ dose administration (t2). Both cohorts present a biased sex ratio (females 79% in HCW and 74% in NHR), as is common in these populations in occident, and together encompass a broad age range (median [age range]: HCW 43 [19-70] and NHR 87 [70-99]). To identify participants with unknown prior infection, the entire HCW cohort was tested at t0 for serum reactivity against SARS-CoV-2 nucleocapsid (N), detecting 26 positive cases (2%). The NHR cohort had no documented previous exposure and all samples collected at day 0 tested negative for anti-spike reactivities. Collected samples were analysed for bulk reactivity against SARS-CoV-2-RBD using a commercial ECLIA (HCW only), and for isotype-specific (IgG, IgM and IgA) anti-SARS-CoV-2-spike using an in-house ELISA assay.

**Figure 1.**
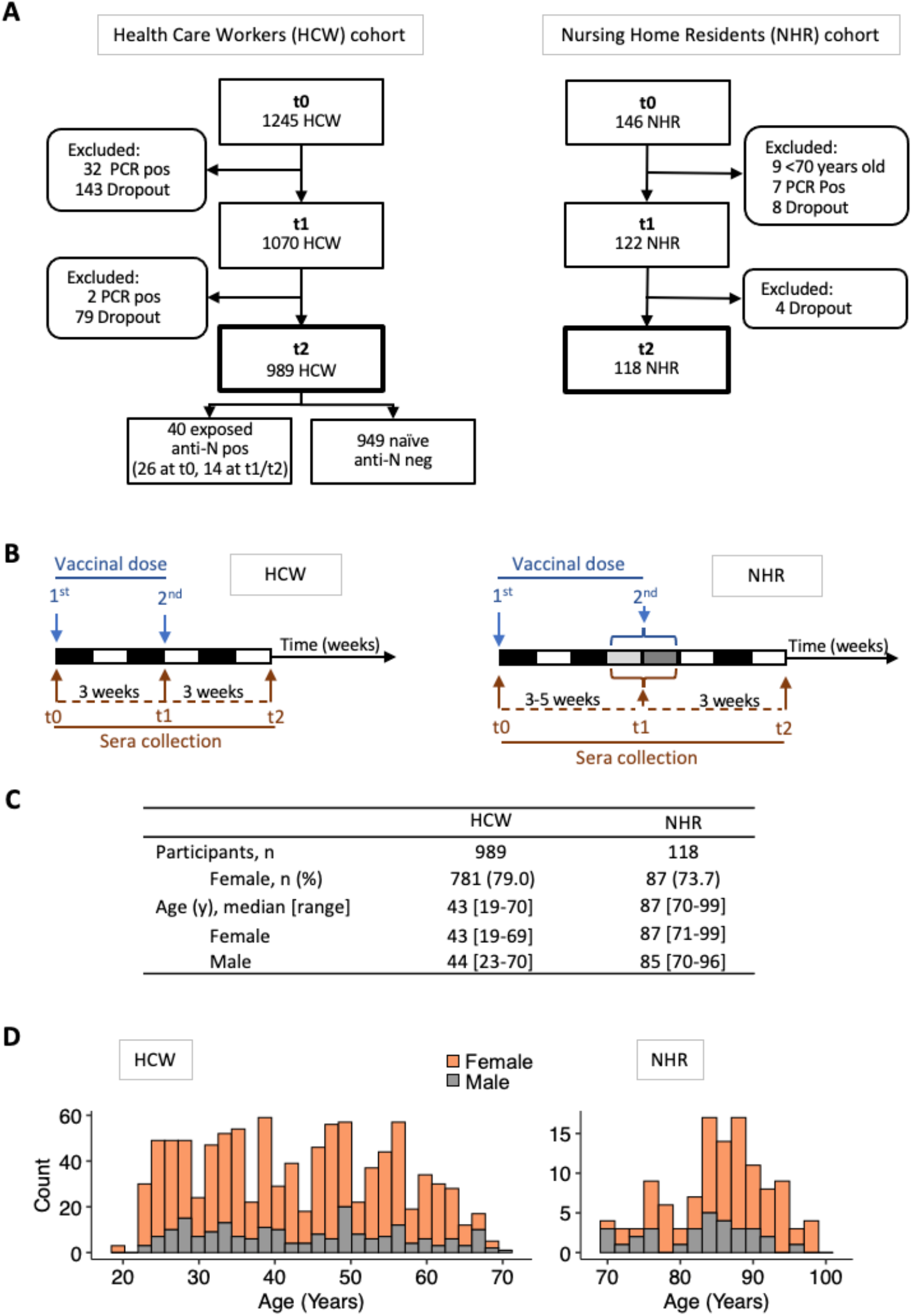
Cohort specification. Hospital healthcare workers (HCW) and nursing home residents (NHR) donated blood samples before vaccination with BNT162b2 RNA (t0), 3-5 weeks after the first dose (t1) and 3 weeks after the second dose (t2). **A**) Enrollment of Participants, exclusion criteria and drop-out. **B**) Collection Schedule. **C**) Age and sex distribution. **D**) stratification by 2-year interval.

### INFECTION

During the course of the study, 41 participants were diagnosed COVID-19 by RT-PCR on nasopharyngeal swab (**Figure 1**). Of these, 39 were infected in the interval between the two vaccine doses (HCW 32/1245, 2.5 % and NHR 7/146, 4,8%, median [IQR] 1.7 [1.14-2.14] weeks post-1^st^ dose for the 2 cohorts). An additional 14 HCW showed *de novo* SARSCoV-2 N antigen reactivity at t1 and/or t2. Diagnosed and inferred cases of COVID-19 post t0 were excluded from the following immunogenicity analysis.

### ISOTYPIC IMMUNOGENICITY

To directly determine the immunogenicity of BNT162b2 vaccine, SARS-CoV-2 naïve participants, defined as negative for serum reactivity anti-SARS-CoV-2 N (HCW, n=949) or spike (NHR, n=118), were first analysed using a binary classification (**Figure 2A**). As expected, seroconversion was the rule in HCW, with bulk anti-RBD reactivity detected at similar frequency whether at 3 weeks post 1^st^ or 2^nd^ injection (99.4% at t1, 99.8 at t2). Isotype class analysis of anti-spike antibodies revealed a heterogeneous response at t1, with 89.2% positivity for IgG, 40.8% for IgM, and 69% for IgA at t1. Increased positivity at t2 was limited to the IgG class, reaching 99.9%. In contrast, seroconversion at t1 was poor in the NHR cohort with only 25.4% positivity for IgG, 2.5 % for IgM and 36.4% for IgA, while the 2^nd^ vaccine dose resulted in 94.9% positivity for IgG reactivities.

Quantitative analysis (**Figure 2B, supplementary Table 1**) revealed very large inter-individual heterogeneity in the amplitude of anti-RBD Ig and anti-spike IgG responses at t1, covering the entire dynamic range of each assays (median [IQR]: HCW 1.44 [1.26-1.57]; NHR 0.50 [0.35-1.06], for anti-S IgG at t1). Large heterogeneity was also observed for anti-spike IgM and IgA levels. Strikingly, the 2^nd^ vaccine dose resulted in major increment and homogenisation to high values of anti-RBD Ig and anti-spike IgG responses, with measurements reaching saturation for the vast majority of participants in HCW as in NHR (median [IQR]: HCW 1.83 [1.72-1.92] and NHR 1.83 [1.68-1.96], for anti-spike IgG at t2). The median anti-spike IgG response was estimated to correspond to titres of at least 1×10^4^ at t2 versus 1×10^3^ and 1×10^2^ at t1 in HCW and NHR, respectively, indicating the 2^nd^ dose provides an increment higher than 10-fold. In contrast, the 2^nd^ vaccine dose does not improve anti-spike-specific IgM and IgA responses, or only mildly as for few NHR participants.

### NON-RESPONDERS

In both cohorts we identified non-responders, defined as not reaching anti-spike IgG level of positivity after 2 vaccine doses (1/949 HCW, 0.1%; 6/118 NHR, 5%). The HCW non-responder, also classified as negative for anti-RBD, was treated for Rheumatoid Arthritis (leflunomide, steroids and methotrexate), and did not respond to a previous Hepatitis B vaccination. The 6 NHR non-responders were 3 males and 3 females of age 84-91 years (median 89), with no apparent association with frailty or medication. Lack of responsiveness in 6 NHR participants, 3 males and 3 females with 84-91 years (median 89), showed no apparent association with frailty or medication.

### CUMULATIVE AGE EFFECT

The parallel analysis of the HCW and NHR cohorts revealed a dramatic effect of advanced age on the magnitude of the antibody response at t1 (**Figure 2**). Stratified analysis by age groups (in 10-year bins) of the HCW cohort revealed an increasing negative effect of age at t1, evident for bulk anti-RBD reactivity and for anti-spike IgG and IgM, but not IgA levels (**Figure 3**). Corroborating this finding, the NHR cohort scored lower than the oldest strata of the HCW (mean [IQR] [age strata]: HCW 1.30 [1.00-1.53] [60-69]; NHR 0.50 [0.35-1.06] [70-99], for anti-spike IgG). Further age stratification of the NHR cohort was not informative.

**Figure 2.**
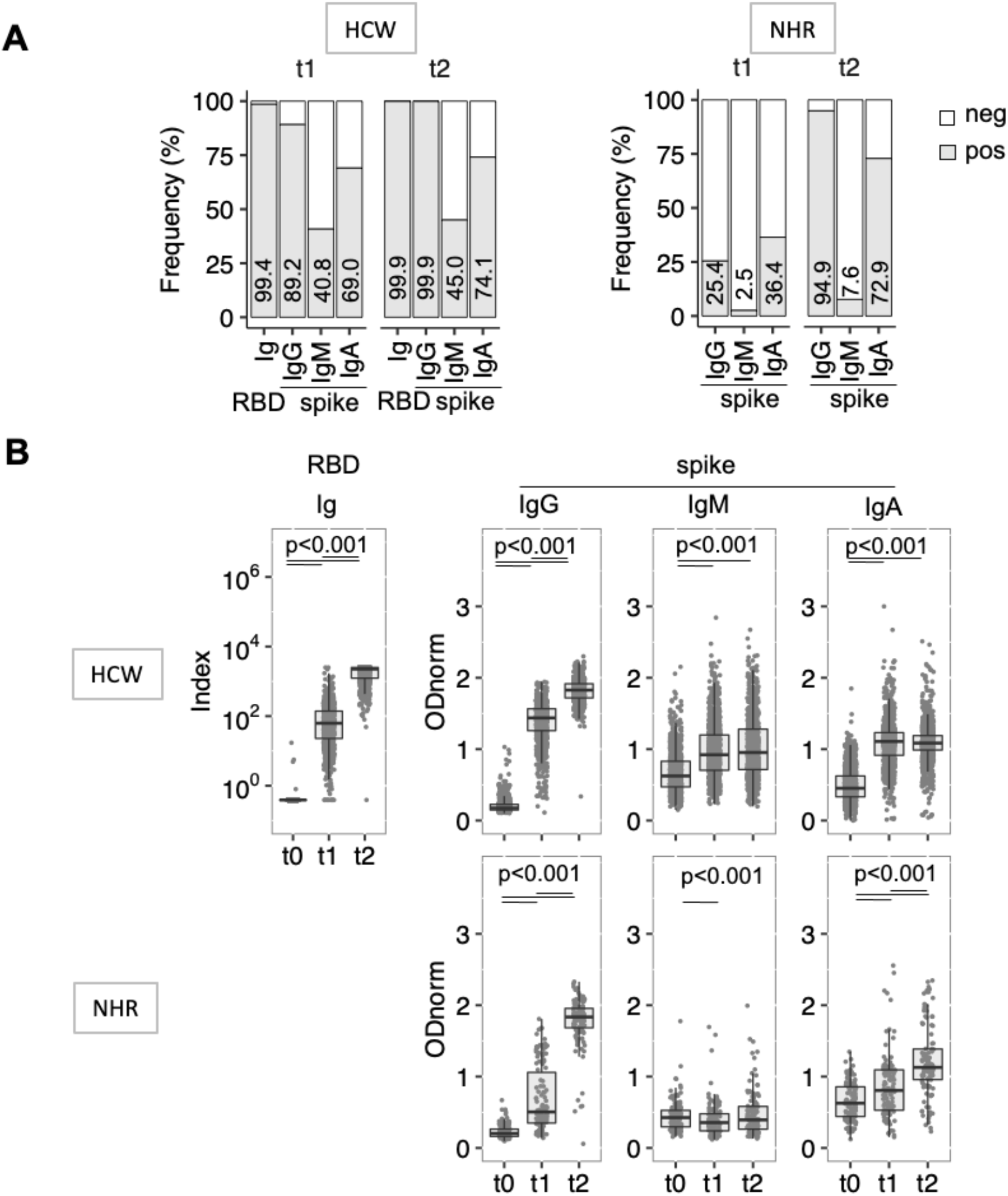
Heterogenous anti-SARS-CoV-2-spike reactivities induced by vaccination. Sera collected as in Fig.1C were analysed for anti-RBD Ig (ECLIA) and anti-full-length spike protein IgG, IgM and IgA (ELISA). Individuals positive for reactivities against SARS-CoV-2 N antigen were removed from the dataset. **A)** Seroconversion defined by frequency of samples testing positive (grey bar) at the indicated day and assays. Respective values are indicated inside each bar. **B)** Semi-quantitative measurements. Data points represent individual participants, boxes denote interquartile range, horizontal line represent the median, and whiskers denote the minimum and maximum values below or above the median at 1.5 times the interquartile range. Note the y scale differs for the anti-RBD ECLIA and the anti-spike ELISA data. Index ≥0,8 and ODnorm≥1 define positivity in (A). Quade test for group differenced across time points, p-value < 0.001 for all panels except NHR IgM where p-value = 0.002. Wilcoxon signed-rank test for pairwise comparison, p-values indicated in figure.

**Figure 3.**
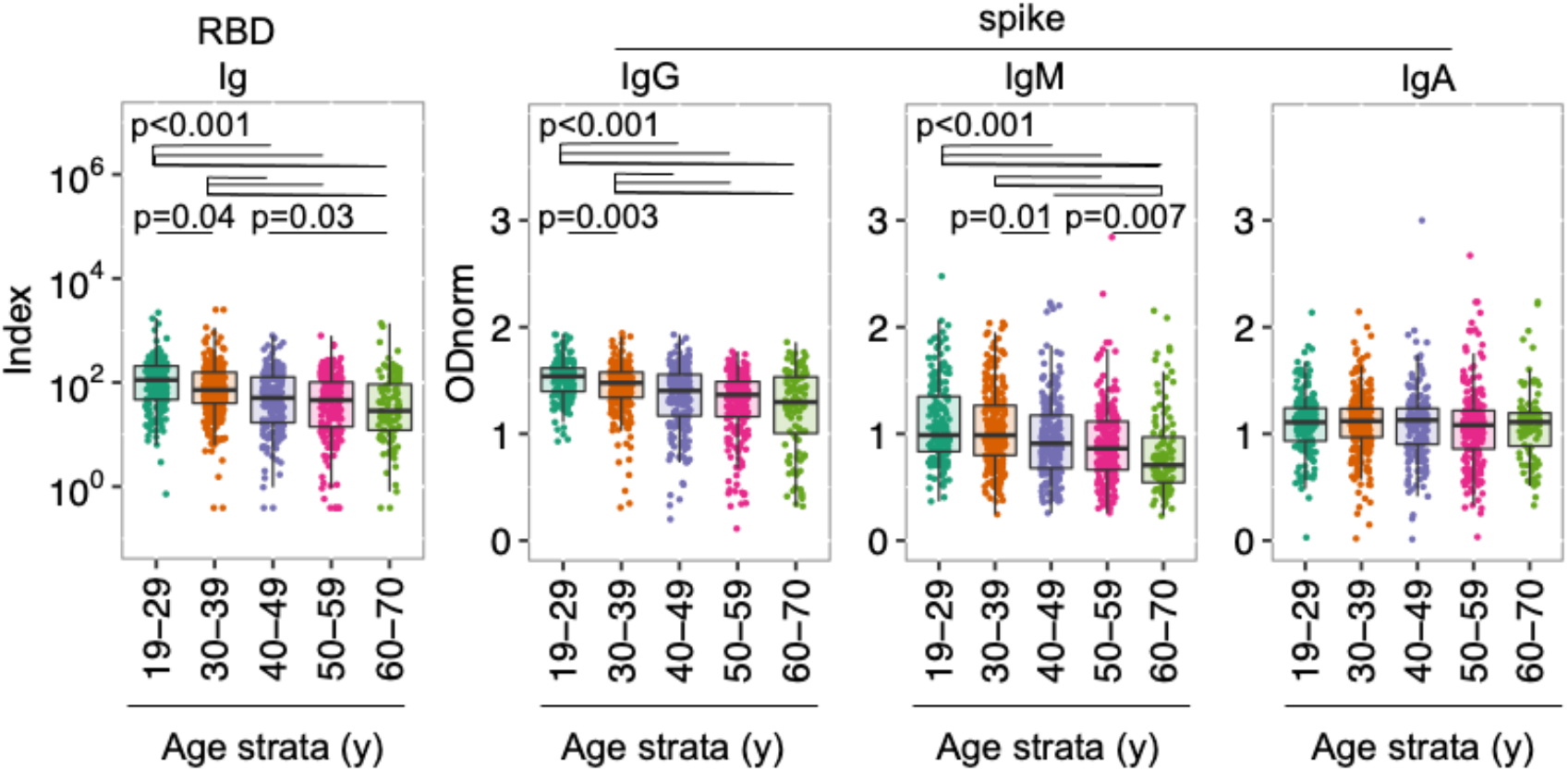
Cumulative effect of age on anti-SARS-CoV-2-spike reactivities induced by vaccination. Shown are semi-quantitative measurements in the HCW cohort at t1, as in Figure 2B, now stratified by age group (interval 10 years). Data points represent individual participants, boxes denote interquartile range, horizontal lines represent the median, and whiskers denote the minimum and maximum values below or above the median at 1.5 times the interquartile range. Kruskal-Wallis test for group difference across age groups, p-value <0.001 for all panels except IgA where p-value=0.55. p-values from Pairwise Wilcoxon rank sum comparison are indicated in each panel.

### SEX EFFECT

Stratification of the cohorts by sex evidenced marginal effect (**Figure 4, Supplementary Table 1 and 2**). After 1^st^ dose administration, males presented lower anti-RBD and anti-spike responses, only in the age stratum 60-69 years, a result that affected also the frequency of IgG seroconversion in this age range (positivity 82.1% for females, 55.2% for males at t1). In the older NHR cohort, IgG levels were marginally higher in females than in males, and only at t2. Despite the overall higher immuno-competence of the youngest age stratum, levels of specific reactivities were still strikingly spread at t1 in this age group (e.g. titre ranges were estimated from 1×10^2^ to 1×10^4^ or higher, for anti-spike IgG in the [19-29] stratum). We excluded concerns of RNA vaccine stability, as stratification by calendar days of the 1^st^ vaccine dose administration showed no effect on antibody levels.

**Figure 4.**
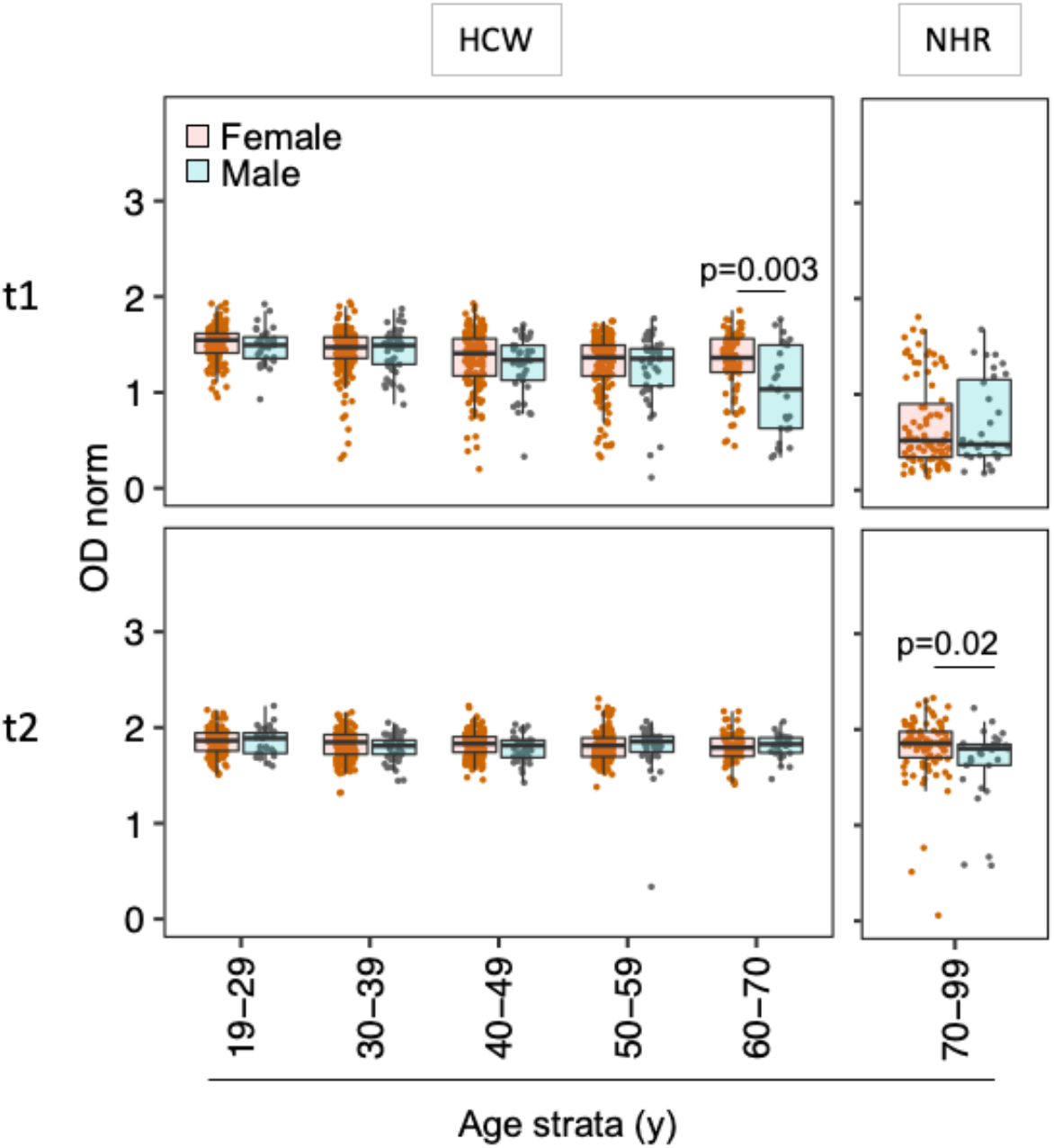
Sex modulates vaccine induced anti-SARS-CoV-2- spike reactivities. **Shown are anti-spike IgG** as in Figure 2B, except that the semi-quantitative measurements are stratified by age and sex (HCW) or sex only (NHR). Data points represent individual females (orange) and males (grey), boxes denote interquartile range, horizontal lines represent the median, and whiskers denote the minimum and maximum values below or above the median at 1.5 times the interquartile range. Kruskal-Wallis tests revealed age group differences in females at t2 (p=0.004), and in males at t1 (p-value <0.001). Wilcoxon rank sum tests at specific age groups revealed sex differences at t1 in HCW aged 60-70 years, and at t2 in NHR, and p-values are indicated in the panels.

### PREVIOUS EXPOSURE

The HCW cohort encompassed 26 participants who tested anti-N positive at t0 (excluded from above analyses). For these participants, anti-RBD Ig and anti-spike IgG levels reached the maximal values of the respective assays by t1 (**Figure 5)**. IgM responses were not significant, while IgA levels reached values slightly above those of naïve participants at t1 (median [IQR]: N-pos 1.51 [1.30 1.85]; N-neg 1.11 [0.91-1.23], at t1), with no enhancement at t2.

**Figure 5.**
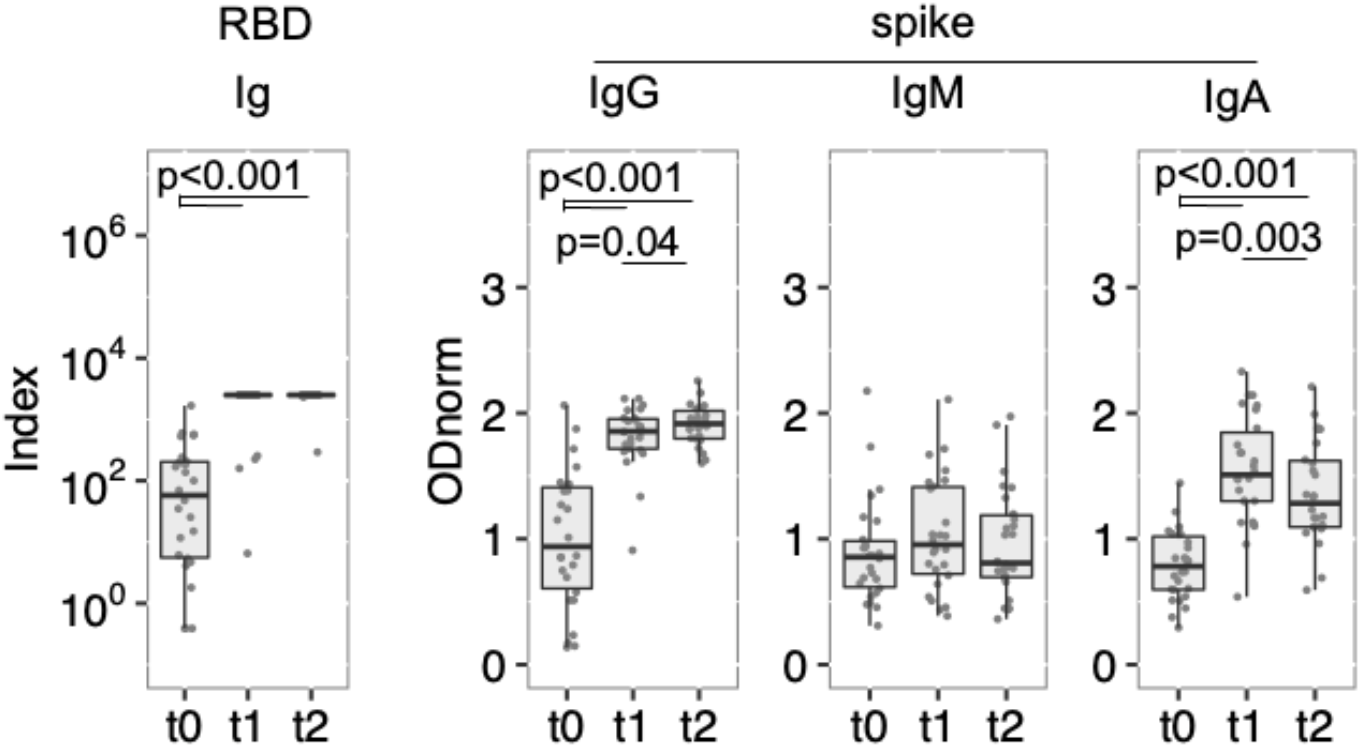
Previous exposure to SARS-CoV-2 enhances vaccine induced anti-SARS-CoV-2-spike reactivities. Shown are semi-quantitative measurement at t0, t1 and t2 for participants identified as anti-N positive at t0, prior to BNT162b2 vaccination. Data points represent individual participants, boxes denote interquartile range, horizontal lines represent the median, and whiskers denote the minimum and maximum values below or above the median at 1.5 times the interquartile range. Quade test for group difference over time points, p-values <0.001 for anti-RBD Ig, anti-spike IgG and IgA, and p=0,24 for anti-spike IgM. Wilcoxon signed-rank test for pairwise comparison, p-values are indicated in each panel.

### DROPOUT

Among the HCW participants who received two vaccine doses and did not develop COVID-19, 222 collected at t0 did not participate in t1 and/or t2 collections (**Figure 1A**). We excluded these would be biased to either low or high reactivities as the median and IQR at t0, t1 and t2 for each isotype were in the range of those of the full cohort (**supplementary Table 3**).

## DISCUSSION

In this study we found a striking inter-individual variation in the amplitude and nature of the humoral response 3-5 weeks after the 1^st^ vaccine dose, explained only in part by age, sex, previous exposure, and drug treatments. These findings have consequences for our understanding of mRNA vaccine immunogenicity, the design of vaccination roll-out and for the management of vaccinees.

Our data indicate BNT162b2 elicits a humoral immune response biased to the IgG class with low contribution of IgM and IgA. This result is consistent with classical IgM responses that peak during the first week post antigen-encounter, and are not significantly boosted through memory cell recall. Of note, our analysis at t0 reveals a sizable fraction of participants presenting IgM anti-spike reactivity prior to vaccination (12.5% above threshold as compared to 0.8% when testing sera from 1000 donors collected before COVID-19 pandemic), and the nature of these peculiar IgM reactivities remains to be understood. Following infection, strong anti-spike IgA responses are frequent, may be more prevalent in symptomatic patients^19^, and confer neutralizing capacity^20^. The rather unchanged anti-spike IgA levels after the 2^nd^ vaccine injection in naïve participants suggests the involvement of T cell independent responses, usually producing monomeric IgA, unlikely to contribute to mucosal immunity upon subsequent SARS-CoV2 infection. Future work will determine whether, as for IgG^21^, specific IgA elicited by mRNA vaccines are present in mucosal secretion.

Identification of a subset of participants testing anti-RBD Ig positive at t0, but unaware of previous exposure to SARS-COV-2, is consistent with a majority of a/pauci-symptomatic cases remaining undetected during 2020. In this subset, the high IgG responses at t1 confirmed the 1^st^ vaccination dose acts as a boost in individuals previously exposed to SARS-CoV-2^7,8,11,12,22^. This finding strengthens the previous proposition that a single vaccine dose may suffice for maximal antibody response in previously infected individuals^13^. In turn, it is plausible that previously exposed non-symptomatic individuals already had reduced specific-antibody levels at t0 but kept immune memory B cells^17^, and these may be the high responders at t1in the group we classified as naïve.

As observed with conventional vaccines^4^, age was a clear factor contributing to decreased amplitude of the response to the 1^st^ dose, though the boosting effect of the 2^nd^ dose was robust in all strata. Sex added an effect to age at t1, in the 60-70 strata only, possibly an indirect effect of behaviour, co-morbidity or treatments^5^.

The limitations of the study include the short follow-up upon the 2^nd^ vaccine dose, and antibody persistence will be addressed in subsequent blood sampling for the same cohorts. The study did not include functional assays such as neutralizing antibodies. However, levels of anti-spike reactivity elicited by BNT162b2 have been previously correlated with *in vitro* neutralization of spike-pseudoviruses and SARS-CoV-2, by ourselves and others^9,23,24^. The study did not include systematic detection of infection cases. Another work addressing a very large population indicated BNT162b2 vaccine effectiveness was of 56.6% after one dose (14-21days) and 96.6% after two doses^25^, confirming the 2^nd^ dose is required for homogeneity at population level. Ad hoc comparison of vaccine effectiveness in this previous report^25^ and seroconversion in our study suggests the threshold of positivity in serological assays maybe revised to higher values to infer protection. More suitable metanalysis will clarify this point.

Those limitations notwithstanding, the low efficacy of single dose BNT162b2 vaccination makes the compelling argument that a second vaccination dose is required to attain uniformly high levels of immunoglobulin in COVID-19 naive individuals. The large spread in the quality of the antibody response 3-5 weeks after the 1^st^ vaccine dose should be taken into account when considering extending the time between first and second administration of BNT162b2 vaccine. This measure was advocated to optimize vaccine roll-out and population protection in the context of limited COVID-19 vaccine supply (e.g. JCVI-UK prolongation for 12 weeks and NACI Canada up to 16 weeks). Corroborating our concern, 32 participants were diagnosed COVID-19 in between the 2 vaccine doses, possibly the combined result of relaxed precaution measures, peak of COVID-19 prevalence, and suboptimal immunity. Similar warning emerged from analysis of cancer patients^24^. Finally, the detection of non-responders by simple reactivity analysis argues for monitoring the post-vaccination antibody level, notably in elderly and immunosuppressed individuals. Serology tests, possibly point of care, upon vaccination and along time, would guide subsequent measures, such as maintaining social distancing but also considering additional vaccine doses and/or switching to other vaccines containing stronger adjuvant components or larger number of epitopes.

## Data Availability

Anonymised raw data for Ig levels will be made available on demand. Biometric data will not be shared.

## Acknowledgements

We thank the healthcare workers and nursing home residents who participated to the study. We are indebted to Jorge Carneiro for help with ELISA data analysis and Tiago Paixão for guidance in data processing. We are grateful to Joao Costa and Joana Bom for providing training on highly specialised equipment made available for this work. We acknowledge the serology4covid consortium for joined effort during the implementation of a low-scale pilot version of the ELISA assay. We also thank the healthcare professionals involved in the HCW sample collection and local testing: Inês Sousa, Catarina Farinha, Susana Vaz, Helena Fernandes, Carla Castro, Catarina Simões, Joana Soares, Nara Silva, Ana Matos, Isabel Barros and Inês Santos. We are indebted to the study assistants who ensured the NHR collection: Lara Fontes, Pureza Duarte Ferreira, Paulo Guia, Pedro Silva and João Ferreira. We are most thankful to all the members of the IGC-COVID19 task force for their continuous support. We thank Antonio Coutinho and Thiago Carvalho for critical reading of the manuscript.

## Supplemental Material

**Supplemental Table 1.**
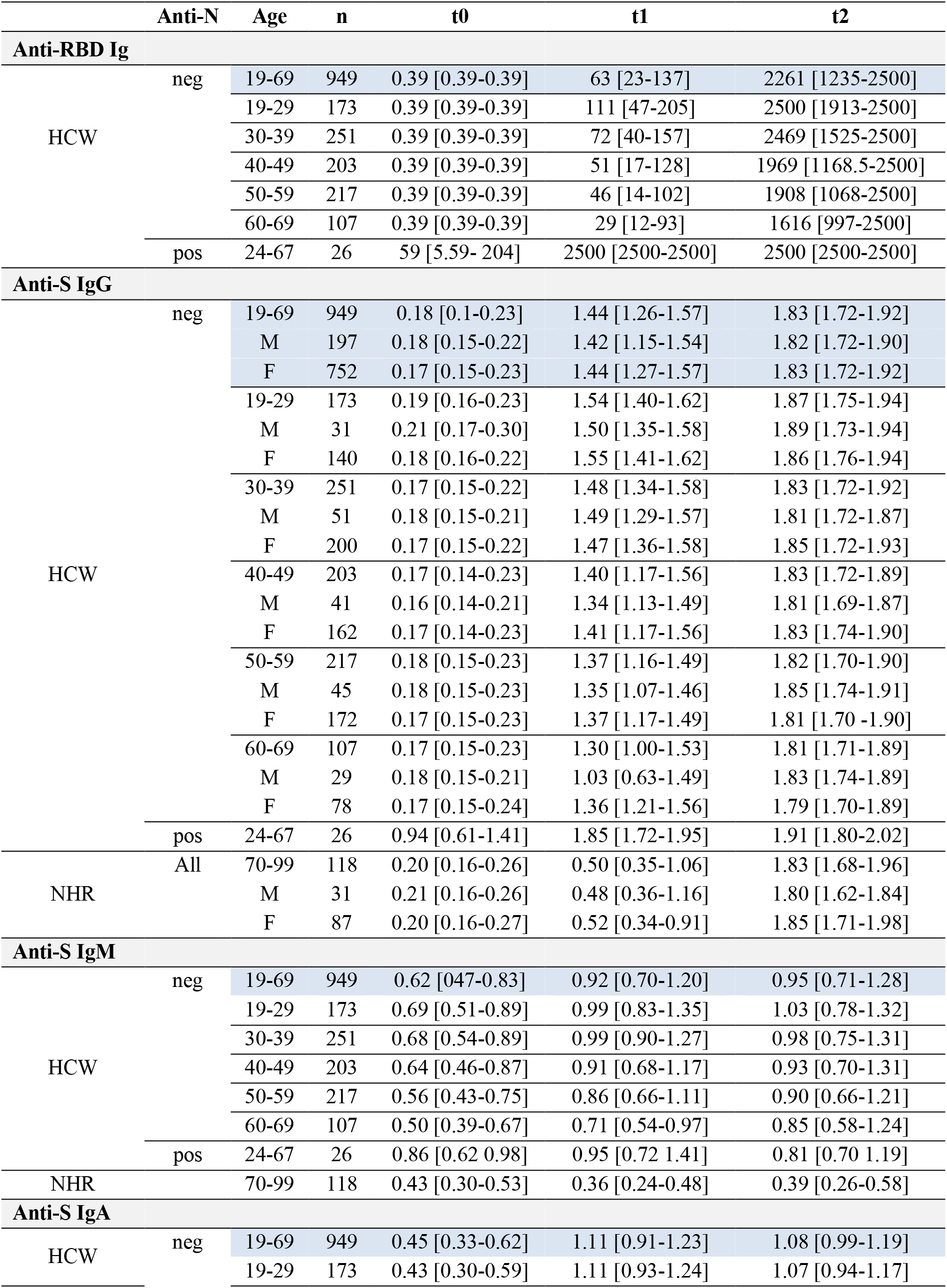
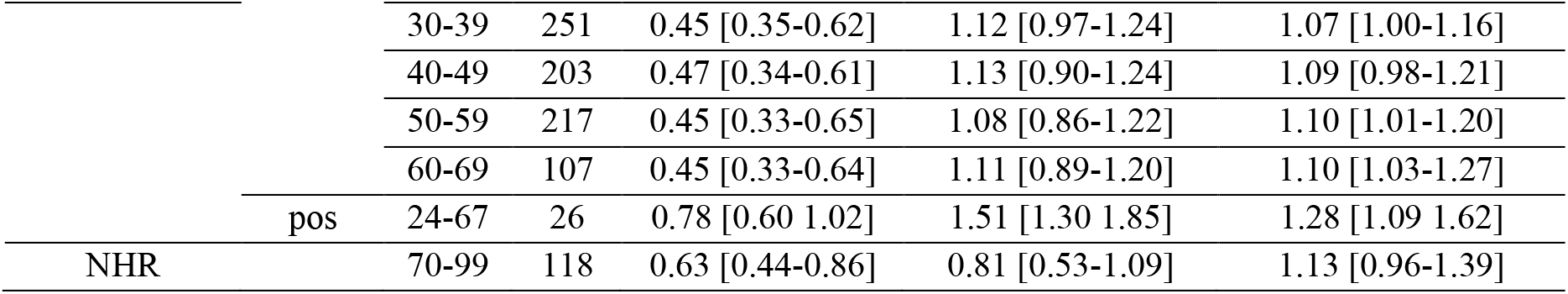
Median and [IQR] for all semi-quantitative data presented in Figures 2-5

**Supplemental Table 2.**
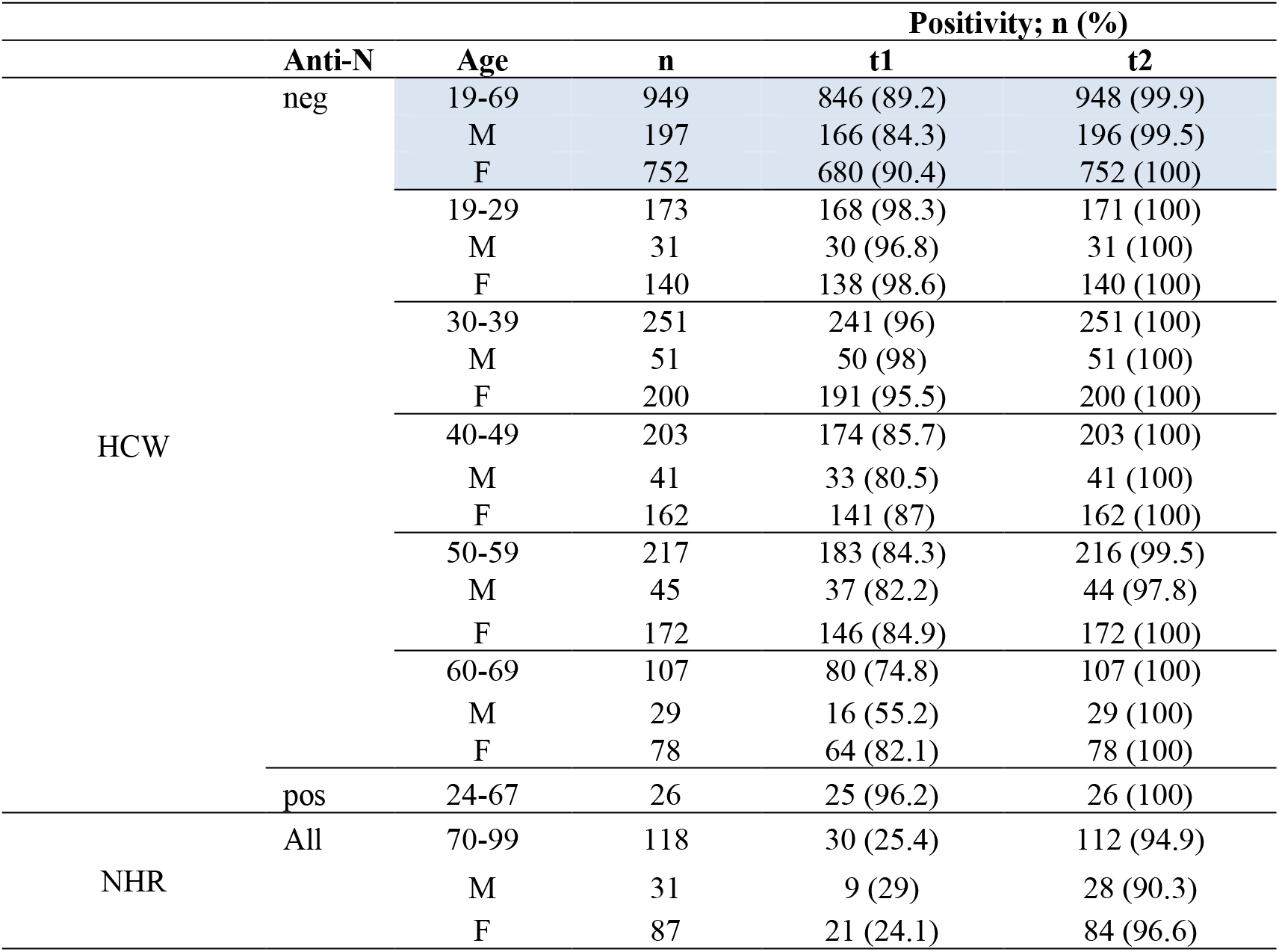
Anti-Spike IgG Positivity in HCW and NHR stratified by age and sex as in Figure 4

**Supplemental Table 3.**
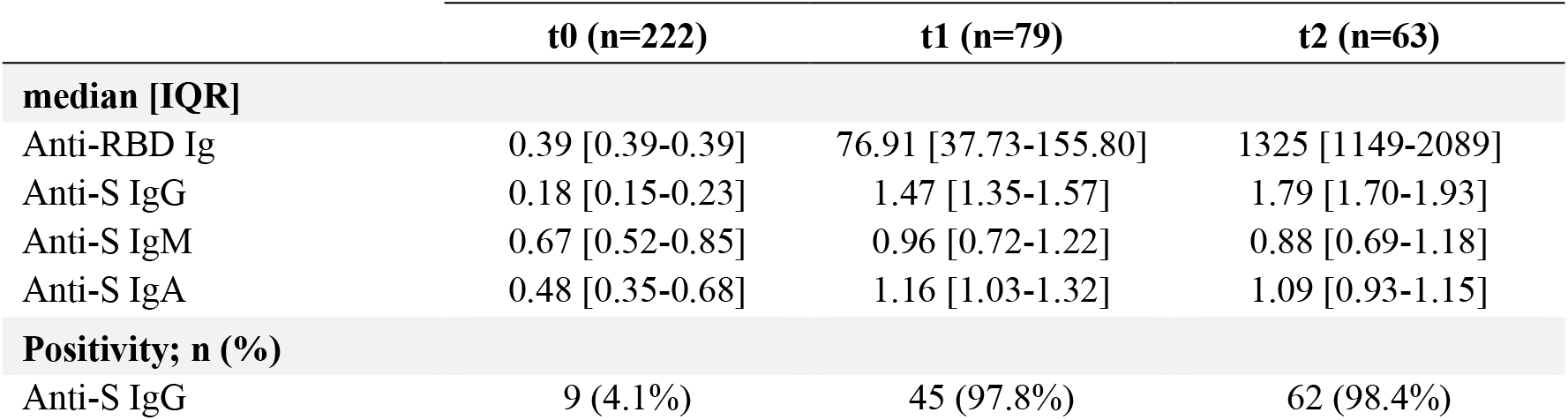
Median and [IQR] for ECLIA and ELISA data, and corresponding anti-spike IgG positivity at each time point, for participants who showed only at one (t0) or two (t0 and t1 or t0 and t2) collection times.

## REFERENCES

1. Polack FP, Thomas SJ, Kitchin N, et al. Safety and Efficacy of the BNT162b2 mRNA Covid-19 Vaccine. N Engl J Med 2020;383:2603–15.

2. Walsh EE, Frenck RW, Jr., Falsey AR, et al. Safety and Immunogenicity of Two RNA-Based Covid-19 Vaccine Candidates. N Engl J Med 2020;383:2439–50.

3. Williamson EJ, Walker AJ, Bhaskaran K, et al. Factors associated with COVID-19-related death using OpenSAFELY. Nature 2020;584:430–6.

4. Ciabattini A, Nardini C, Santoro F, Garagnani P, Franceschi C, Medaglini D. Vaccination in the elderly: The challenge of immune changes with aging. Semin Immunol 2018;40:83–94.

5. Klein SL, Flanagan KL. Sex differences in immune responses. Nat Rev Immunol 2016;16:626–38.

6. Wang Z, Schmidt F, Weisblum Y, et al. mRNA vaccine-elicited antibodies to SARS-CoV-2 and circulating variants. Nature 2021.

7. Manisty C, Otter AD, Treibel TA, et al. Antibody response to first BNT162b2 dose in previously SARS-CoV-2-infected individuals. Lancet 2021.

8. Krammer F, Srivastava K, Alshammary H, et al. Antibody Responses in Seropositive Persons after a Single Dose of SARS-CoV-2 mRNA Vaccine. N Engl J Med 2021.

9. Doria-Rose N, Suthar MS, Makowski M, et al. Antibody Persistence through 6 Months after the Second Dose of mRNA-1273 Vaccine for Covid-19. New England Journal of Medicine 2021.

10. Anderson EJ, Rouphael NG, Widge AT, et al. Safety and Immunogenicity of SARS-CoV-2 mRNA-1273 Vaccine in Older Adults. New England Journal of Medicine 2020;383:2427–38.

11. Saadat S, Rikhtegaran Tehrani Z, Logue J, et al. Binding and Neutralization Antibody Titers After a Single Vaccine Dose in Health Care Workers Previously Infected With SARS-CoV-2. JAMA 2021;325:1467–9.

12. Anichini G, Terrosi C, Gandolfo C, et al. SARS-CoV-2 Antibody Response in Persons with Past Natural Infection. New England Journal of Medicine 2021.

13. Wise J. Covid-19: People who have had infection might only need one dose of mRNA vaccine. BMJ 2021;372:308.

14. Liu Y, Liu J, Xia H, et al. Neutralizing Activity of BNT162b2-Elicited Serum. New England Journal of Medicine 2021;384:1466–8.

15. Ju B, Zhang Q, Ge J, et al. Human neutralizing antibodies elicited by SARS-CoV-2 infection. Nature 2020;584:115–9.

16. Sattler A, Angermair S, Stockmann H, et al. SARS-CoV-2-specific T cell responses and correlations with COVID-19 patient predisposition. J Clin Invest 2020;130:6477–89.

17. Sherina N, Piralla A, Du L, et al. Persistence of SARS-CoV-2-specific B and T cell responses in convalescent COVID-19 patients 6-8 months after the infection. Med (N Y) 2021;2:281–95 e4.

18. Amanat F, Stadlbauer D, Strohmeier S, et al. A serological assay to detect SARS-CoV-2 seroconversion in humans. Nat Med 2020;26:1033–6.

19. Sterlin D, Mathian A, Miyara M, et al. IgA dominates the early neutralizing antibody response to SARS-CoV-2. Sci Transl Med 2021;13.

20. Wang Z, Lorenzi JCC, Muecksch F, et al. Enhanced SARS-CoV-2 neutralization by dimeric IgA. Sci Transl Med 2021;13.

21. Mades A, Chellamathu P, Lopez L, et al. Detection of persistent SARS-CoV-2 IgG antibodies in oral mucosal fluid and upper respiratory tract specimens following COVID-19 mRNA vaccination. medRxiv 2021.2021.05.06.21256403.

22. Ebinger JE, Fert-Bober J, Printsev I, et al. Antibody responses to the BNT162b2 mRNA vaccine in individuals previously infected with SARS-CoV-2. Nature Medicine 2021.

23. Alenquer M, Ferreira F, Lousa D, et al. Amino acids 484 and 494 of SARS-CoV-2 spike are hotspots of immune evasion affecting antibody but not ACE2 binding. bioRxiv 2021:2021.04.22.441007.

24. Monin L, Laing AG, Muñoz-Ruiz M, et al. Safety and immunogenicity of one versus two doses of the COVID-19 vaccine BNT162b2 for patients with cancer: interim analysis of a prospective observational study. The Lancet Oncology 2021.

25. Haas EJ, Angulo FJ, McLaughlin JM, et al. Impact and effectiveness of mRNA BNT162b2 vaccine against SARS-CoV-2 infections and COVID-19 cases, hospitalisations, and deaths following a nationwide vaccination campaign in Israel: an observational study using national surveillance data. The Lancet 2021;397:1819–29.

